# Plasma Hydrogen Sulfide Production Capacity is Positively Associated with Post-Operative Survival in Patients Undergoing Surgical Revascularization

**DOI:** 10.1101/2021.02.16.21251804

**Authors:** Alban Longchamp, Michael R. MacArthur, Kaspar Trocha, Janine Ganahl, Charlotte G. Mann, Peter Kip, William W. King, Gaurav Sharma, Ming Tao, Sarah J. Mitchell, Tamás Ditrói, Péter Nagy, C. Keith Ozaki, Christopher Hine, James R. Mitchell

## Abstract

**Objective:** Hydrogen sulfide (H_2_S) is a gaseous signaling molecule and redox factor important for cardiovascular function. Deficiencies in its production or bioavailability are implicated in atherosclerotic disease. However, it is unknown if circulating H_2_S levels differ between vasculopaths and healthy individuals, and if so, whether H_2_S measurements can be used to predict surgical outcomes. Here, we examine: 1) Plasma H_2_S levels in patients undergoing vascular surgery and compare these to healthy controls, and 2) Associations between H_2_S levels and mortality in surgical revascularization patients.

**Approach & Results:** Patients undergoing carotid endarterectomy, open lower extremity revascularization or leg amputation were enrolled. Peripheral blood was also collected from a matched cohort of 20 patients without peripheral or coronary artery disease. Plasma H_2_S production capacity and sulfide concentration were measured using the lead acetate and monobromobimane methods, respectively. Plasma H_2_S production capacity and plasma sulfide concentrations were reduced in patients with PAD (p<0.001, p=0.013 respectively). Patients that underwent surgical revascularization were divided into high versus low H_2_S production capacity groups by median split. Patients in the low H_2_S production group had increased probability of mortality (p=0.003). This association was robust to correction for potentially confounding variables using Cox proportional hazard models.

**Conclusions:** Circulating H_2_S levels were lower in patients with atherosclerotic disease. Patients undergoing surgical revascularization with lower H_2_S production capacity, but not sulfide concentrations, had increased probability of mortality within 36 months post-surgery. This work provides insight on the role H_2_S plays as a diagnostic and potential therapeutic for cardiovascular disease.

**HIGHLIGHTS:**

- Vascular disease patients have higher plasma hydrogen sulfide levels than controls without vascular disease as measured by two distinct methods, the lead acetate hydrogen sulfide release method and the HPLC-based monobromobimane method.
- Only the lead acetate hydrogen sulfide release method robustly predicts survival after vascular surgery intervention over 35 months of follow up.
- The lead acetate release method measures non-enzymatic hydrogen sulfide release from plasma which requires iron and is catalyzed by vitamin B_6_.

## INTRODUCTION

Hydrogen sulfide (H_2_S) is a redox modifying and diffusible gasotransmitter that plays numerous physiologic roles across various organ systems including the cardiovascular system^1^. While high levels of exogenous H_2_S are toxic, increased levels within a supraphysiologic range have been shown to mediate many beneficial effects ranging from stress resistance to longevity^2-4^. H_2_S has also emerged as a critical mediator of vascular homeostasis^5^ through its functions as a vasodilator^6^, antioxidant^7^, oxygen sensor^8, 9^, immunomodulator^10^, and anti-inflammatory gas^11^. Importantly, as atherosclerosis is a chronic inflammatory disease, it is important to note H_2_S is shown to regulate several atherosclerotic cellular and inflammatory processes^12-15^.

While H_2_S has been implicated in the pathogenesis of multiple cardiovascular disease processes, the measurement of H_2_S is arduous and often relies on indirect measures and surrogates^16-21^. Nevertheless, characterizing the role of H_2_S in systemic disease states such as peripheral arterial disease (PAD) may yield important insights on potential diagnostic and therapeutic applications.

We therefore assessed H_2_S levels in human plasma using two measurement methods; the lead acetate H_2_S production capacity quantification method and the monobromobimane method (MBB method) for sulfide quantification. Patients included in this study were diagnosed with either carotid artery stenosis requiring carotid endarterectomy (CEA), or peripheral artery disease (PAD) necessitating revascularization or amputation secondary to unsalvageable critical limb ischemia. Patients were prospectively followed for 36 months post-surgery with clinical outcomes and mortality measured. A baseline control cohort of patients matched for age, sex, and hypertension^6^, but with no histories of PAD, prior MI, coronary interventions, heart failure, or stroke were included for comparing H_2_S levels in the diseased versus healthy state. Our study provides new insight into the relevance of circulating H_2_S levels in patients suffering from atherosclerosis, and during surgical revascularization.

## MATERIAL AND METHODS

### Study Design & Demographic Data Collection

Patients at a single institution undergoing elective vascular surgery including patients scheduled for carotid endarterectomy, open lower extremity revascularization, and major lower extremity amputation were enrolled. Informed consent was obtained for prospective collection of demographic and clinical data under a Partners Human Research Committee institutional review board approved protocol.

Surgical patients enrolled encompassed patients with carotid artery stenosis requiring CEA for asymptomatic (>80% stenosis) or symptomatic (>50% stenosis) carotid disease, patients with PAD requiring infra-inguinal surgical revascularization and patients with secondary critical limb ischemia (CLI) refractory to lifestyle and surgical interventions requiring major lower extremity amputation (above knee or below knee amputation).

Baseline patient risk factors and demographics were collected including age, race, history of PAD, history of stroke or myocardial infarction as well as history of coronary intervention (**Supplemental Table 1**). To compare levels of H_2_S between patients with vascular disease vs. healthy patients, a group of 20 control subjects were enrolled without documented or diagnosed PAD or CAD. This control cohort were matched for age, sex and hypertension. Patients were excluded if they were <18 years, had an emergent indication for the operation or if they were involved in another clinical research study.

### Hydrogen Sulfide Measurements

On the day of surgery, but prior to surgical intervention, peripheral venous blood was collected into ethylenediaminetetraacetic acid (EDTA) collection tubes at the time of peripheral intravenous line placement. The tube was inverted several times to ensure mixing with the anticoagulant and then transferred to the lab for processing. To obtain plasma all samples were centrifuged at room temperature for 15 minutes at 2,000g, plasma was then stored at −80°C for future analysis. All samples were then thawed and processed for H_2_S production capacity at once to ensure uniformity.

Hydrogen sulfide production capacity was measured using the lead acetate method^22^. In brief, plasma was mixed with 150ul freshly prepared reaction mixture, containing 100 mM L-cysteine and 0.5 mM PLP in Phosphate buffered saline (PBS) in a plastic 96-well plate. The plate was then incubated at 37°C with lead acetate embedded filter paper on top. Upon the reaction of H_2_S with the lead acetate paper, a dark lead sulfide precipitate is produced. The paper was incubated until a detectable, but non-saturated signal was seen. The amount of lead sulfide captured on the paper was quantitated by measuring the integrated density of each well area using ImageJ software.

Hydrogen sulfide concentrations using the MBB method was conducted as described previously^23^. Briefly: In almost complete darkness due to the light sensitivity of the MBB reagent, 20 µl of 50 mM HEPES (pH=8.0) buffer and 20 µl of 10mM monobromo-bimane (Sigma Aldrich) was added to 20 µl of plasma sample. After 10 minutes in the dark at room temperature, the product sulfide-dibimane was extracted with 200 µl of pure ethyl acetate. The organic supernatant was collected and evaporated to dryness and stored at −20°C until measurement. The solid samples were redissolved prior to HPLC measurements in acetonitrile. 10 µl was injected and separated on an Agilent Eclipse XDB-C18 (4.6×250mm, 5µm) using a Merck Hitachi L7000 HPLC instrument with a Thermo UltiMate 3000 fluorescent detector. The elution method employed a 28 min long gradient using water and acetonitrile both containing 0.1% TFA. The detection of the product was carried out using UV-absorbance measurement at 254 nm and fluorescent measurement with extinction at 390 nm and detection at 480 nm. Quantitation was done using a standard calibration curve in aqueous buffered solutions, where H_2_S concentrations were verified by the DTNB assays. It should be noted that the MBB method measures endogenous sulfide levels that are easily liberated from the bound plasma sulfide pool and hence absolute values largely depend on the applied conditions (temperature, alkylation time, concentration conditions etc). This is the reason why absolute values should not be compared with values reported in other studies^21^.

The enzymatic activity of CBS in blood samples was measured by an HPLC-MS/MS protocol previously published^24^. Briefly, 20 μL of plasma sample was added to 25 μL of solution containing 200 mmol/L Tris-HCl (pH 8.6), 1 mmol/L pyridoxal 5′-phosphate, 1 mmol/L SAM and 40 mmol/L 2,3,3-2H-labeled serine (Cambridge Isotope Laboratories, Inc). 5 μL of starting solution containing 280 mmol/L homocysteine thiolactone in 100 mmol/L Tris-HCl (pH 8.6), 10 mmol/L DTT, 1.225 mol/L NaOH was incubated for 5 minutes at 37°C to produce homocysteine by thiolactone cleavage, pH was adjusted to 8.6 with 1:1 HCl solution and the solution was added to the plasma mixture. After 4 hours of incubation at 37°C, the reaction was quenched by acidification of the mixture with 100 μL of EZ:faast Internal Standard Solution (Phenomenex) containing 3.3 μmol/L of internal standard 3,3,4,4-2H-labeled cystathionine. Sample preparation was carried out using EZ:faast kit (Phenomenex) kit, that included a solid phase extraction step, derivatization with propyl chloroformate, and an extraction into an organic solvent. The prepared samples were separated on an EZ:faast AAA-MS column (250 × 2.0 mm, Phenomenex) using LC and MS settings described in the EZ:faast user manual using a Thermo Scientific UltiMate 3000 HPLC connected to a Thermo Scientific LTQ-XL MS instrument. Concentrations (and enzyme activities) were calculated using the internal standard.

### Statistics

Plasma H_2_S production capacity and sulfide concentration in vascular surgery patients and healthy controls were compared using Student’s t test. Pearson correlation coefficient was calculated to assess the association between plasma H_2_S production capacity and plasma sulfide concentration. The Kaplan-Meier method was used to estimate survival in high and low H_2_S production capacity/free sulfide groups and groups were compared using log-rank test. Cox proportional hazard models were fit to estimate mortality hazard ratios between high and low H_2_S producing groups, corrected for potentially confounding variables (age, gender, BMI). Both unadjusted and adjusted Cox proportional hazard models met assumptions of proportional hazard. To assess the odds of post-surgical complication in high vs low H_2_S producing individuals, Fisher’s exact test was used. Statistical tests were performed using Graphpad Prism 7 and R version 3.3.2. Kaplan-Meier curves and Cox proportional hazard models were fit in R using the Survival package and Kaplan-Meier curves were visualized using the Survminer package. All reported P values are based on 2-sided tests and P values of less than 0.05 were considered statistically significant.

## RESULTS

Results using the lead acetate-based H_2_S production capacity assay showed that healthy controls had significantly higher H_2_S production capacity compared to vascular disease patients (**Fig1A, S1A**, 64.4±5.1 vs 40.16±1.8 arbitrary units, p < 0.001). Results using the MBB method also showed that healthy controls had significantly greater plasma sulfide levels (**Fig1B**, 0.95±0.07 vs 0.76±0.03 µM, p < 0.05). Although both of these sulfide-based measurements were significantly higher in healthy controls, there was no observable correlation between the measurements (**Fig1C**, r_s_ = −0.12, p = 0.34), suggesting that the two methods measure fundamentally different phenomena.

**Figure 1.**
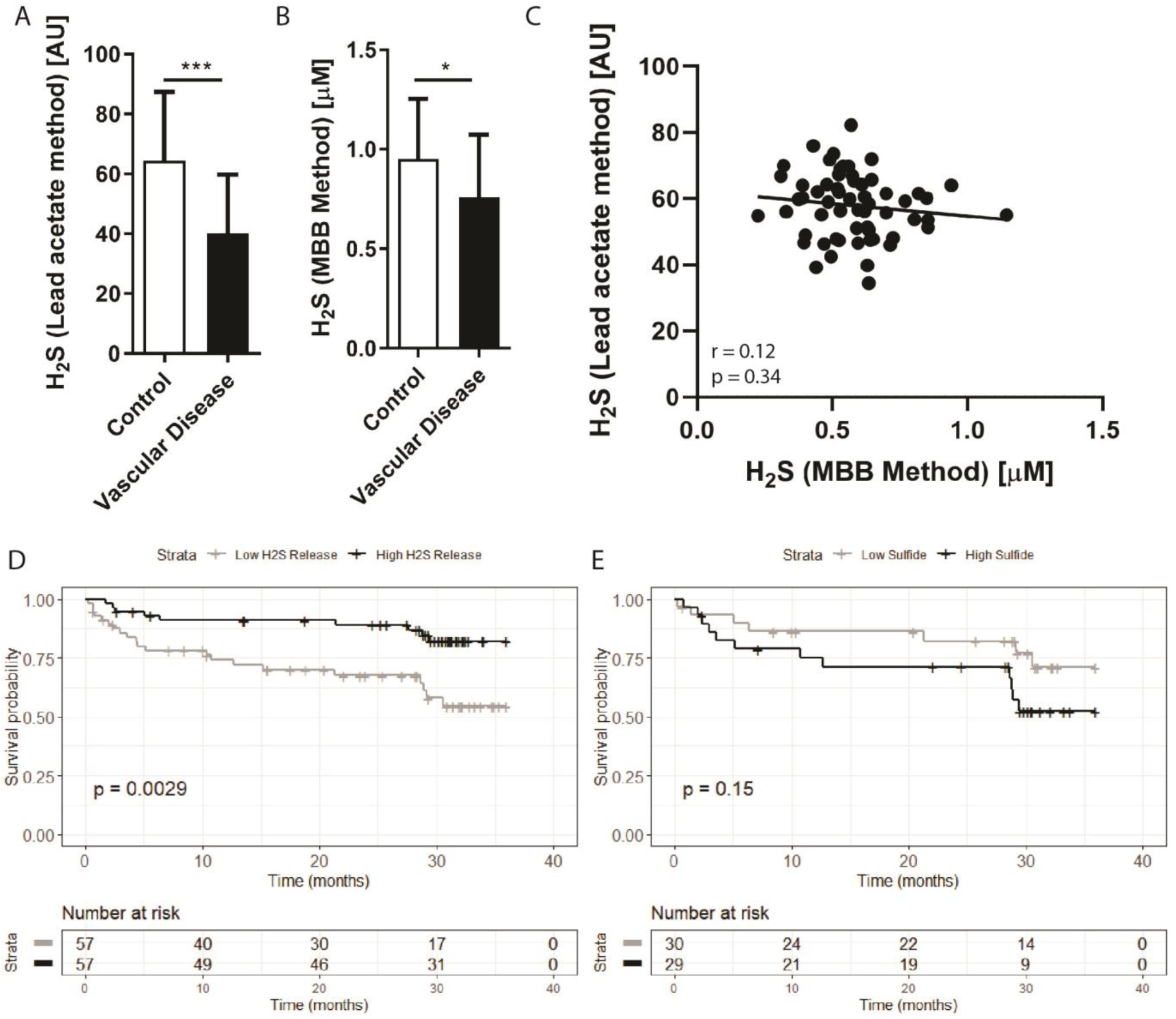
Plasma H_2_S production capacity and sulfide levels are reduced in patients with vascular disease, with production capacity predicting mortality. (**A**) Plasma H_2_S production capacity and **(B)** plasma sulfide measured by the MBB method from human patients suffering vascular occlusive disease (n=115) and healthy age-matched individuals (n=20). Error bars indicate SD; * p<0.05, ***p<0.001. (**C**) Correlation between H_2_S production capacity and sulfide measured by the MBB method in vascular disease patients. **(D)** Probability of survival for vascular disease patients during follow up after intervention with low (n=57) versus high (n=57) H_2_S production capacity and **(E)** low and high plasma sulfide measurements. High versus low was determined by median split. P value calculated from log-rank test.

To assess the associations of these H_2_S measurements with clinical outcomes after surgical procedures, the vascular surgery patients were divided by median split into patients with high and low pre-operative H_2_S production capacity (**FigS1B**). Over 36 months of follow up, patients with low H_2_S production capacity had significantly decreased probability of survival compared to patients with high H_2_S production capacity (**Fig1D**, 54% vs 82% survival probability, log-rank test p = 0.0029). However, when patients were divided by median split into high and low MBB-measured sulfide measurements, there was no significant association with survival and the trend was reversed such that those with higher sulfide tended to have lower probability of survival (**Fig1E**, 71% vs 52% survival probability, log-rank test = 0.15).

To further assess the robustness of H_2_S production capacity as a predictor of mortality after a vascular surgery intervention, we generated Cox proportional hazard models to adjust for potential confounding variables. A univariate unadjusted model showed that individuals in the high H_2_S production capacity group had significantly reduced risk of death during follow up (HR(95% CI) = 0.31(0.11 – 0.87), coefficient p=0.025, model Wald p = 0.025). In a Cox proportional hazard model adjusted for age, BMI, gender, smoking history and race, H_2_S production capacity was significantly associated with survival (HR=0.94, CI=0.90-0.99, coefficient p=0.017, model Wald p = 0.009, **Figure 2**).

**Figure 2.**
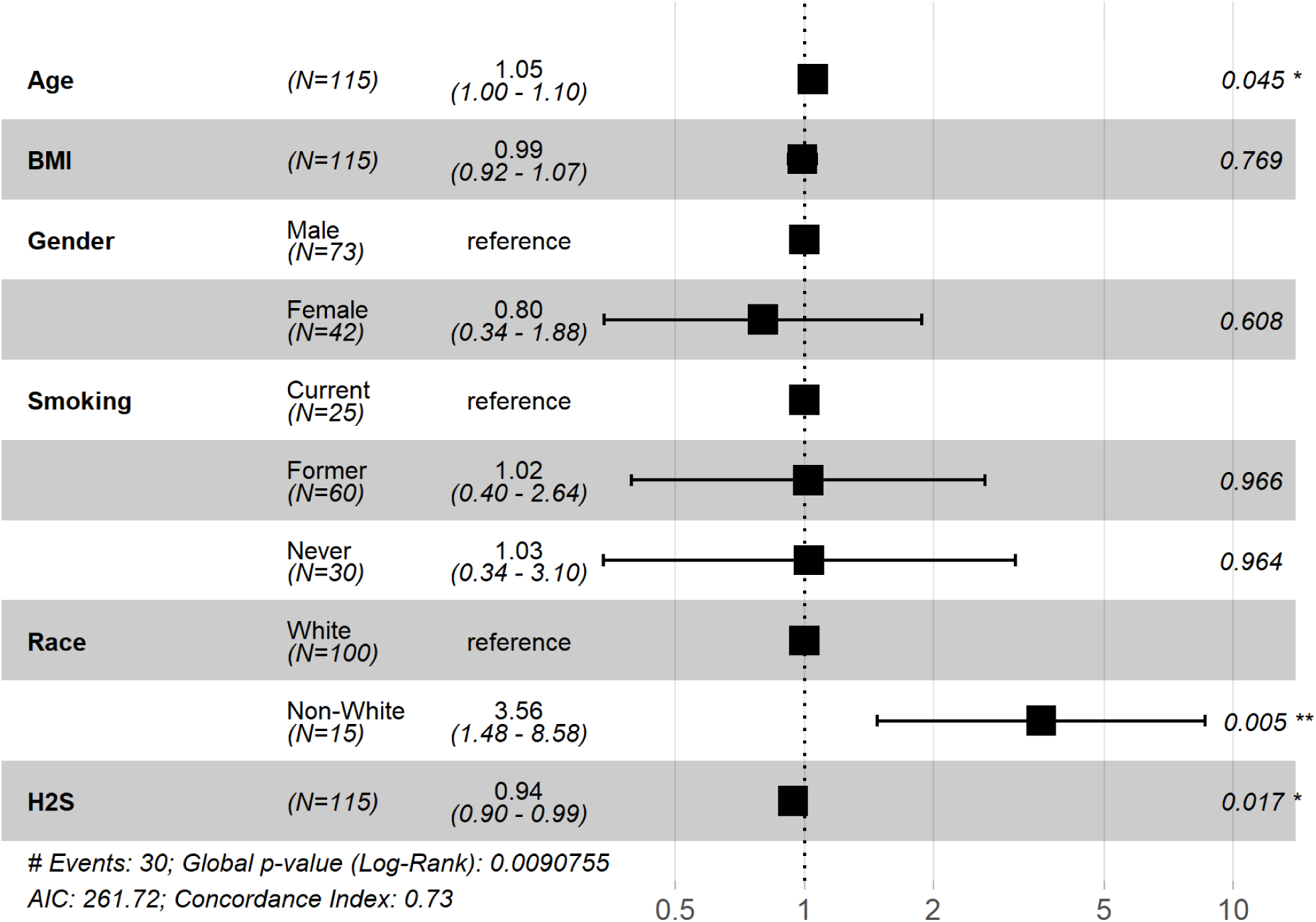
Hazard ratios of death during follow up period. Hazard ratio estimates and 95% confidence intervals for clinically relevant parameters and plasma H_2_S production capacity as measured by the lead acetate release method

Finally, to characterize the biochemical nature of the plasma H_2_S production capacity measurement, we investigated whether it might be generated by enzymatic or non-enzymatic production. It has been proposed that the H_2_S generating enzyme cystathionine beta synthase (CBS) is present in plasma. We measured plasma CBS activities, but found no observable association between plasma CBS activity and H_2_S production capacity (**FigS1C**, r = −0.07, p = 0.71). We next performed the assay under denaturing conditions by including heating and adding DTT and found a highly correlated pattern of signals as when performed under non-denaturing conditions (**FigS1D**, r = 0.34, p = 0.0002). These results suggest H_2_S production arising from a non-enzymatic process. Finally, we added chelators and found that EDTA was able to almost completely ablate the H_2_S release, while EGTA was not (**FigS1E**). On the other hand, EDTA had a reverse effect on measured sulfide concentrations by the MBB method^21^ corroborating that the two methods measure sulfide released from fundamentally different endogenous pools. This suggests a vital role for iron as a catalyst of the measured H_2_S release. These results on the probably non-enzymatic nature of the signal are in agreement with the recent thorough characterization of mechanisms of H_2_S release in blood by Hine and colleagues^25^.

## DISCUSSION

Pre-clinical studies have suggested that decreased levels of H_2_S accelerate the development of atherosclerosis^12^, and are reduced in the skeletal muscle of CLI patients^1^. Here we show that patients with vascular disease have significantly decreased circulating H_2_S production capacity and sulfide concentrations, compared to subjects with no clinical evidence of coronary or peripheral artery disease. Together, these results suggest that patients with atherosclerotic vascular disease have a decreased capability to generate H_2_S. Furthermore, patients with higher H_2_S production capacity measured prior to vascular surgery had reduced post-operative mortality at 36 months follow-up compared to those with lower H_2_S production. Interestingly, this association was not observed with sulfide levels that were measured by the MBB-method. While this suggest that although both measures are related to sulfide release from bound sulfide reserves, they specifically capture distinct biological phenomena with clinical relevance. Endogenous enzymatic biosynthesis of H_2_S in cells relies on three pyridoxal-5-phosphate (PLP) dependent enzymes responsible for the metabolism of the amino acid L-cysteine. These enzymes being cystathionine-gamma-lyase (CGL), cystathionine-beta-synthase (CBS)^26^ and cysteinyl t-RNA synthetase (CARS2)^27^ as well as a non-PLP dependent enzyme, 3-mercaptopyruvate sulfurtransferase (3-MST)^28^ which are responsible for much H_2_S production in numerous cell types including endothelial cells. Here we show that the clinically relevant H_2_S production capacity measurement captures non-enzymatic H_2_S production.

Hine and colleagues have recently performed a thorough chemical characterization of the mechanisms of non-enzymatic H_2_S production in blood, demonstrating that it is catalyzed by iron and vitamin B_6_ (PLP)^25^. Although this work gives us many clues, important questions remain about what specific components of the plasma determine and regulate H_2_S production capacity. One theory revolves around the sulfur containing amino acid homocysteine, which in itself has been long associated with cardiovascular disease risk, but its mechanism of pathology is not well understood^29^. Homocysteine has been reported to capture H_2_S and/or HS^-^ to form a homocysteine persulfide in cardiac tissue, which could potentially interfere with H_2_S-related cardiovascular protection^30^. Thus, if surgical patients with decreased survival showed an increase in plasma homocysteine levels, then this could explain the dichotomy in H_2_S release in these patients using the two different methods (lead acetate vs. MBB), as homocysteine can trap H_2_S under non-denaturing conditions in the MBB method.

Furthermore, in the non-enzymatic production of H_2_S in plasma catalyzed by iron described previously^25^ and confirmed here, PLP still acts as an important co-factor by the formation of a Schiff base and subsequent cysteine-aldimine and thiazolidine five-member ring intermediates prior to iron rapidly catalyzing the release of the sulfide. Importantly, homocysteine itself can also form a Schiff base with PLP, except it will result in a more thermodynamically stable six-member tetrahydrothiazine ring^31^. The formation of the stable tetrahydrothiazine ring poses two complications for H_2_S production in circulation: the first being sequestration of valuable PLP co-factor from both enzymatic and non-enzymatic H_2_S production where cysteine serves as substrate; and the second being slow H_2_S production kinetics when iron serves as a catalyst^25^.

In addition to addressing the issues brought up in the discussion, the results reported here will aid in the development of specific H_2_S assays for diagnostic purposes. Further, they will guide therapeutic interventions if these specific determinants are shown to be causal to the disease process. Despite these remaining questions, we demonstrate that the lead acetate assay represents a simple, rapid and very low-cost method which appears to capture substantial information on clinical risk in this population.

Limitations of our study include the quantification of H_2_S using the lead acetate method measures only relative differences in H_2_S between individuals and groups, and not absolute differences. Likewise, it primarily serves as a surrogate for the actual amount of H_2_S produced from available substrates in plasma. It is also important to note that mortality outcomes in this study encompassed two uniquely different patient populations, patients with carotid stenosis and patients with PAD necessitating surgical revascularization or amputation. These differences may influence disparities in mortality outcomes and future work will need to validate these findings in larger cohorts with more homogenous interventions. Nevertheless, we highlight our important results showing that both H_2_S production capacity and MBB-method-measured sulfide levels in vascular patients were significantly reduced compared to healthy subjects, indicating a potential correlation between H_2_S and the progression of cardiovascular pathology. These results provide further insights into the role of H_2_S biology in surgical patients and open an avenue for the use of H_2_S for diagnostics and therapeutics in those with dysfunction of their vascular system.

## Data Availability

Data may be provided to qualified researchers upon request

## NONSTANDARD ABREVIATIONS AND ACRONYMS

CARS2: Cysteinyl t-RNA synthetase
CBS: Cystathionine-beta-synthase
CEA: Carotid endarterectomy
CGL: Cystathionine-gamma-lyase
CLI: Critical limb ischemia
H_2_S: Hydrogen sulfide
MI: Myocardial infarction
MBB: Monobromobimane
PAD: Peripheral arterial disease
PLP: Pyridoxal-5-phosphate

## ACKNOWLEDGMENT

none

---

**Figure S1.**
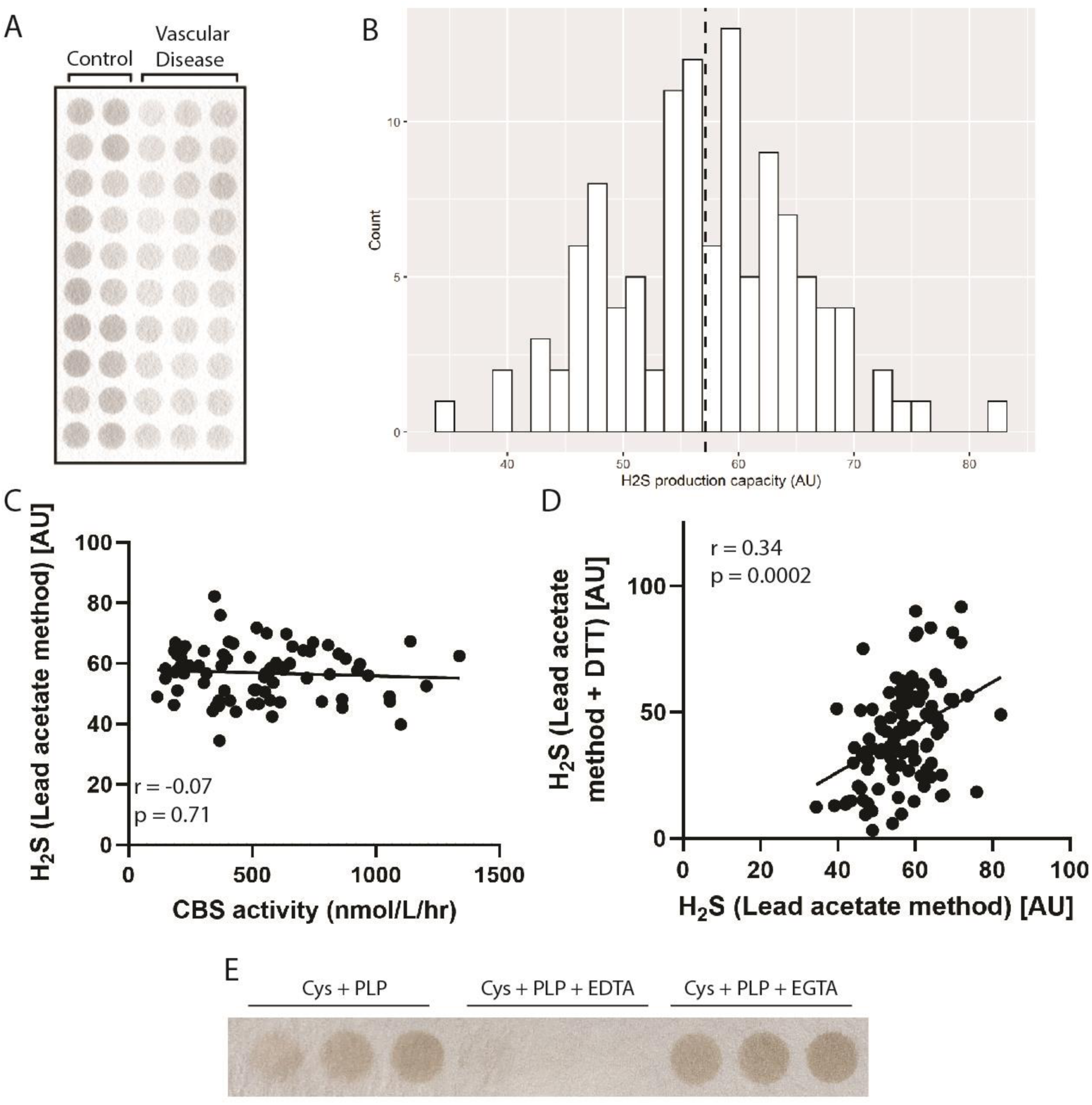
Plasma H_2_S production capacity occurs by a non-enzymatic, iron-dependent process. **(A)** Image of H_2_S released from plasma onto lead acetate paper. **(B)** Histogram of quantified lead acetate H_2_S release from healthy and diseased individuals. **(C)** Correlation between plasma H_2_S production capacity and plasma cystathionine beta synthase protein activity. **(D)**Correlation between plasma H_2_S production capacity and plasma H_2_S production capacity under denaturing conditions with DTT. **(E)** Plasma H_2_S production capacity under normal assay conditions (cysteine plus PLP added), or with added EDTA (12.5mM) or EGTA (12.5mM

**Supplemental Table 1:** Baseline Study Population Characteristics

| *Variable* | *Control Cohort n=20* | *Vascular Cohort n=115* |
| --- | --- | --- |
| Age (SD) | 68 (2.3) | 69(9) |
| Male (%) | 13 (65) | 73 (63) |
| Ethnicity (%) |  |  |
| White | 18 (90) | 97 (84) |
| Hispanic | 0 | 10 (9) |
| Black | 0 | 4 (3.5) |
| Other | 2 (10) | 4 (3.5) |
| BMI (SD) | 28.7 (7.2) | 28.3 (5.3) |
| Comorbidities (%) |  |  |
| Heart failure | 0 | 19 (17) |
| History of stroke | 0 | 24 (21) |
| PAD | 0 | 75 (65) |
| Prior coronary intervention | 0 | 44 (38) |
| Prior MI | 0 | 30 (26) |
| Hypertension | 19 (95) | 106 (92) |
| Hyperlipidemia | 6 (30) | 98 (85) |
| COPD | 0 | 7 (6) |
| Renal dysfunction | 5 (1) | 16 (14) |
| Diabetes mellitus | 2 (10) | 45 (39) |
| Smoking status (%) |  |  |
| Never | 9 (45) | 25 (22) |
| Former | 11 (55) | 60 (52) |
| Current | 0 | 30 (26) |
| Renal dysfunction | 5 (1) | 16 (14) |
| Diabetes mellitus | 2 (10) | 45 (39) |
| Procedure type (%) |  |  |
| Carotid endarterectomy | 0 | 49 (43) |
| Infragaunal revascularization | 0 | 44 (38) |
| Lower extremity amputation | 0 | 22 (19) |

SD, standard deviation; BMI, body mass index; MI, myocardial infarct; HTN, hypertension; COPD, chronic obstructive pulmonary disease

